# Polygenic background modifies penetrance of monogenic variants conferring risk for coronary artery disease, breast cancer, or colorectal cancer

**DOI:** 10.1101/19013086

**Authors:** Akl C. Fahed, Minxian Wang, Julian R. Homburger, Aniruddh P. Patel, Alexander G. Bick, Cynthia L. Neben, Carmen Lai, Deanna Brockman, Anthony Philippakis, Patrick T. Ellinor, Christopher A. Cassa, Matthew Lebo, Kenney Ng, Eric S. Lander, Alicia Y. Zhou, Sekar Kathiresan, Amit V. Khera

## Abstract

**Background:** Genetic variation can predispose to disease both through (i) monogenic risk variants in specific genes that disrupt a specific physiologic pathway and have a large effect on disease risk and (ii) polygenic risk that involves large numbers of variants of small effect that affect many different pathways. Few studies have explored the interaction between monogenic risk variants and polygenic risk.

**Methods:** We identified monogenic risk variants and calculated polygenic scores for three diseases, coronary artery disease, breast cancer, and colorectal cancer, in three study populations — case-control cohorts for coronary artery disease (UK Biobank; N=12,879) and breast cancer (Color Genomics; N=19,264), and an independent cohort of 49,738 additional UK Biobank participants.

**Results:** In the coronary artery disease case-control cohort, increased risk for carriers of a monogenic variant ranged from 1.3-fold for those in the lowest polygenic score quintile to 12.6-fold for those in the highest. For breast cancer, increased risk ranged from 2.4 to 6.9-fold across polygenic score quintiles. Among the 49,738 UK Biobank participants who carried a monogenic risk variant, the probability of disease at age 75 years was strongly modified by polygenic risk. Across individuals in the lowest to highest percentiles of polygenic risk, the probability of disease ranged from 17% to 78% for coronary artery disease; 13% to 76% for breast cancer; and 11% to 80% for colon cancer.

**Conclusions:** For three important genomic conditions, polygenic risk powerfully modifies the risk conferred by monogenic risk variants.

## Background

For a range of common heritable diseases, a small subset of the population inherits a rare monogenic variant that causes a large increase in disease risk by disrupting a specific physiological pathway. More recently, polygenic scores have been developed that integrate the effects of many common genetic variants on disease risk. While the common variants have small individual effects (in the range of a few percent) on disease risk, they can cumulatively have large effects—producing, in some individuals, risks equivalent to the strong monogenic variants.^1–4^

A key open question is how monogenic and polygenic risk interact: Can disease risk from a monogenic variant that causes major disruption to a specific pathway be meaningfully modified by polygenic risk factors that involve small perturbations to a wide range of cellular pathways? Taking familial hypercholesterolemia as an example, monogenic variants predispose to premature coronary artery disease through major dysregulation of clearance of LDL cholesterol from the circulation. By contrast, only a small minority (∼20%) of common DNA variants that predispose to coronary artery disease operate via cholesterol-related pathways, with the remainder affecting non-cholesterol related pathways (such as inflammation, cellular proliferation, vascular tone) and many additional pathways yet to be discovered.^5^ Similarly, pathway analyses indicate that less than 20% of common variants linked to breast or colorectal cancer affect genes involved in DNA repair pathways, which are implicated in monogenic hereditary breast and ovarian cancer or Lynch syndrome.^3,6^

We explored this question by performing genetic analysis for three diseases—familial hypercholesterolemia, hereditary breast and ovarian cancer, and Lynch syndrome predisposing to colorectal cancer—for which about 1% of the population inherits a monogenic variant having a strong effect on disease risk.^7,8^

Although such variants are associated with several-fold increased risk of disease, it has long been recognized that the variants have incomplete penetrance and variable expressivity. For example, in one U.S. healthcare system, more than 50% of familial hypercholesterolemia variant carriers and more than 70% of female hereditary breast and ovarian cancer variant carriers remained free of disease well into middle age.^9,10^

Polygenic risk might help explain variation in outcomes for carriers of monogenic variants. If so, it could have important fundamental implications both for understanding disease physiology and for genetic counseling.

Here, we studied in 81,881 individuals to examine whether polygenic risk can account for variation in outcomes for carriers of monogenic variants—a topic that has both scientific implications about disease physiology and clinical implications for genetic counseling.

## Methods

### Study populations

We studied three diseases: coronary artery disease, breast cancer, and colorectal cancer. For coronary artery disease, we designed and studied a case-control cohort (6,449 cases and 6,430 controls) derived from the UK Biobank, a prospective cohort study that enrolled middle-aged adult participants between 2006 and 2010.^11^ We followed up these results in an independent cohort of 49,738 participants of the UK Biobank who previously underwent exome sequencing.^12^ For breast cancer, we studied an all female case-control cohort (1,920 cases and 17,344 controls) from a commercial testing laboratory (Color Genomics; Burlingame, CA), and then followed up the results in 27,144 female UK Biobank participants. For colorectal cancer, we studied 49,738 UK Biobank participants.

For participants in the UK Biobank, cases of coronary artery disease, breast cancer, and colorectal cancer were identified based on a combination of self-reported data confirmed by trained healthcare professionals, hospitalization records, and national procedural, cancer, and death registries. For participants in the commercial testing cohort, breast cancer status was defined based on self-report at the time of enrollment.^13^ Additional details on study cohorts, genetic testing, and disease ascertainment are provided in the Supplementary Appendix.

### Monogenic variant classification

For each of three conditions, observed variants were classified as pathogenic or likely pathogenic by a laboratory geneticist according to current clinical standards blinded to any phenotype data.^14^ We analyzed three genes causal for familial hypercholesterolemia (*LDLR, APOB*, and *PCSK9*), two for hereditary breast and ovarian cancer syndrome (*BRCA1* and *BRCA2*), and four for Lynch syndrome (*MLH1, MSH2, MSH6*, and *PMS2*).

### Polygenic background

To quantify the contribution of polygenic background to disease, we calculated previously validated polygenic scores for each of the three diseases.^1–3^ Ancestry-corrected reference distributions for each score were generated, as described previously (Fig. S1 in the Supplementary Appendix).^15^

### Statistical Analysis

In the coronary artery disease and breast cancer case-control studies, logistic regression was used to determine risk among participants stratified by monogenic variant status and polygenic score. Within the cohort of 49,738 UK Biobank participants, we modeled the odds ratio for disease according to monogenic variant status and polygenic score percentiles. A second analysis estimated probability of disease by age 75 years using Cox regression, standardized to the mean of all covariates in each population. All regression models were adjusted for age, sex, and genetic ancestry — as quantified by the first four principal components — except for breast cancer analyses, which were restricted to females.^16^

## Results

### Polygenic background modifies risk of coronary artery disease conferred by familial hypercholesterolemia variants

We first studied the interaction of monogenic risk variants and polygenic scores in coronary artery disease. To identify individuals with monogenic variants causal for familial hypercholesterolemia, we sequenced the three genes related to the condition — *LDLR, APOB*, and *PCSK9* — in 6,449 coronary artery disease cases and 6,430 controls derived from the UK Biobank (Table 1). Each of the observed genetic variants was reviewed by a laboratory geneticist blinded to any phenotype data. A total of 28 distinct genetic variants were classified as pathogenic or likely pathogenic; they were present in 43 (0.67%) cases and 13 (0.20%) controls. The presence of a familial hypercholesterolemia variant conferred a 3.17-fold increased risk of coronary artery disease (95% confidence interval [CI] 1.70 to 5.92).

**Table 1:**
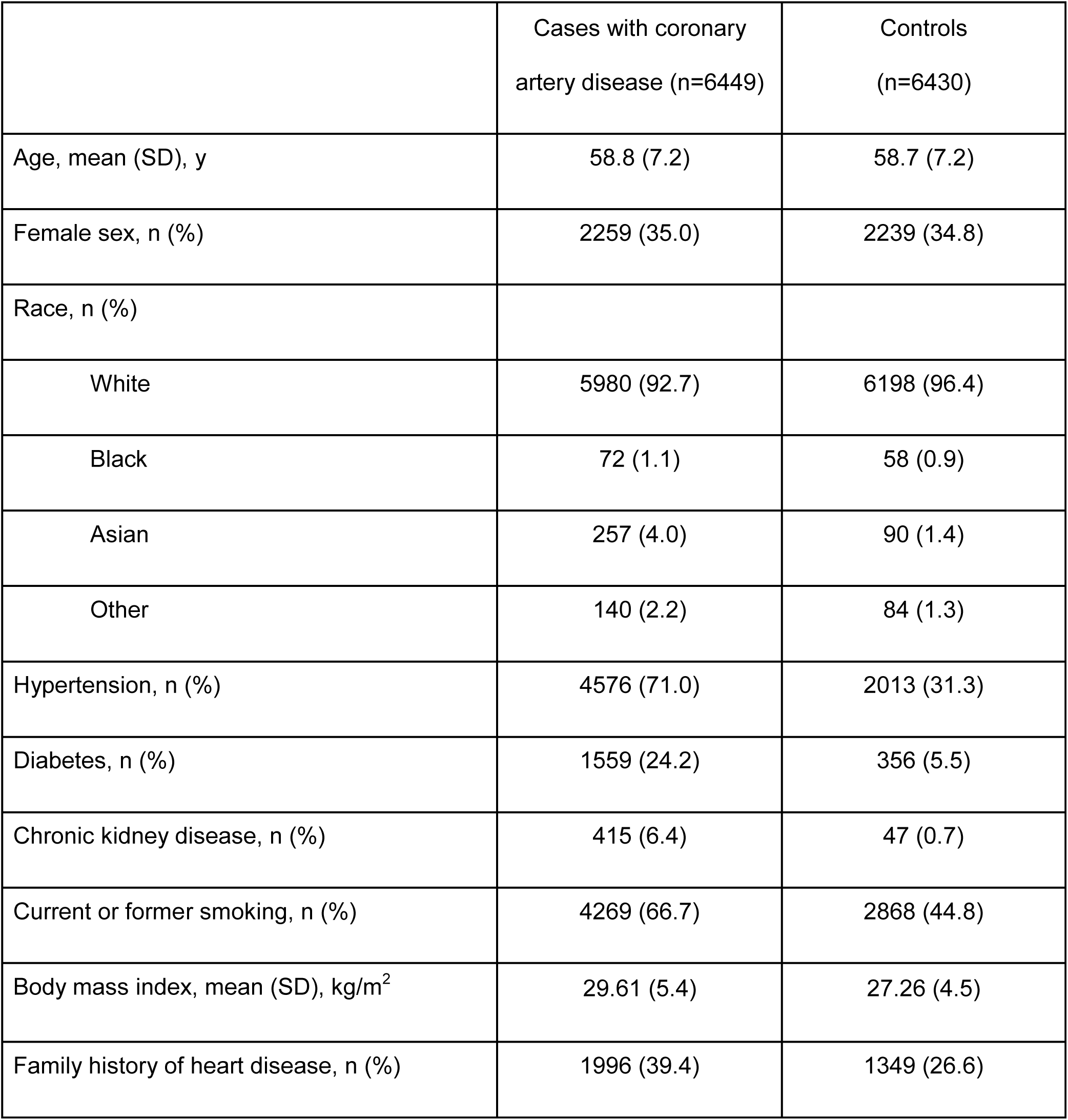
Baseline characteristics of coronary artery disease case-control study participants derived from the UK Biobank

We next examined the effect of participants’ polygenic background on risk of coronary artery disease, by computing a previously validated polygenic score in all cases and controls. Even among carriers of a familial hypercholesterolemia variant, the observed risk varied substantially according to the polygenic score. We classified individuals as having low polygenic score (bottom quintile), intermediate polygenic score (quintiles 2-4), or high polygenic score (top quintile). Compared to individuals who did not carry a pathogenic mutation, the risk among mutation carriers ranged from 1.30-fold (95% CI 0.39 to 4.31) for those in the lowest quintile of the polygenic score distribution to 12.59 (95% CI 2.96 to 53.53) in the highest quintile (Figure 1A).

**Figure 1:**
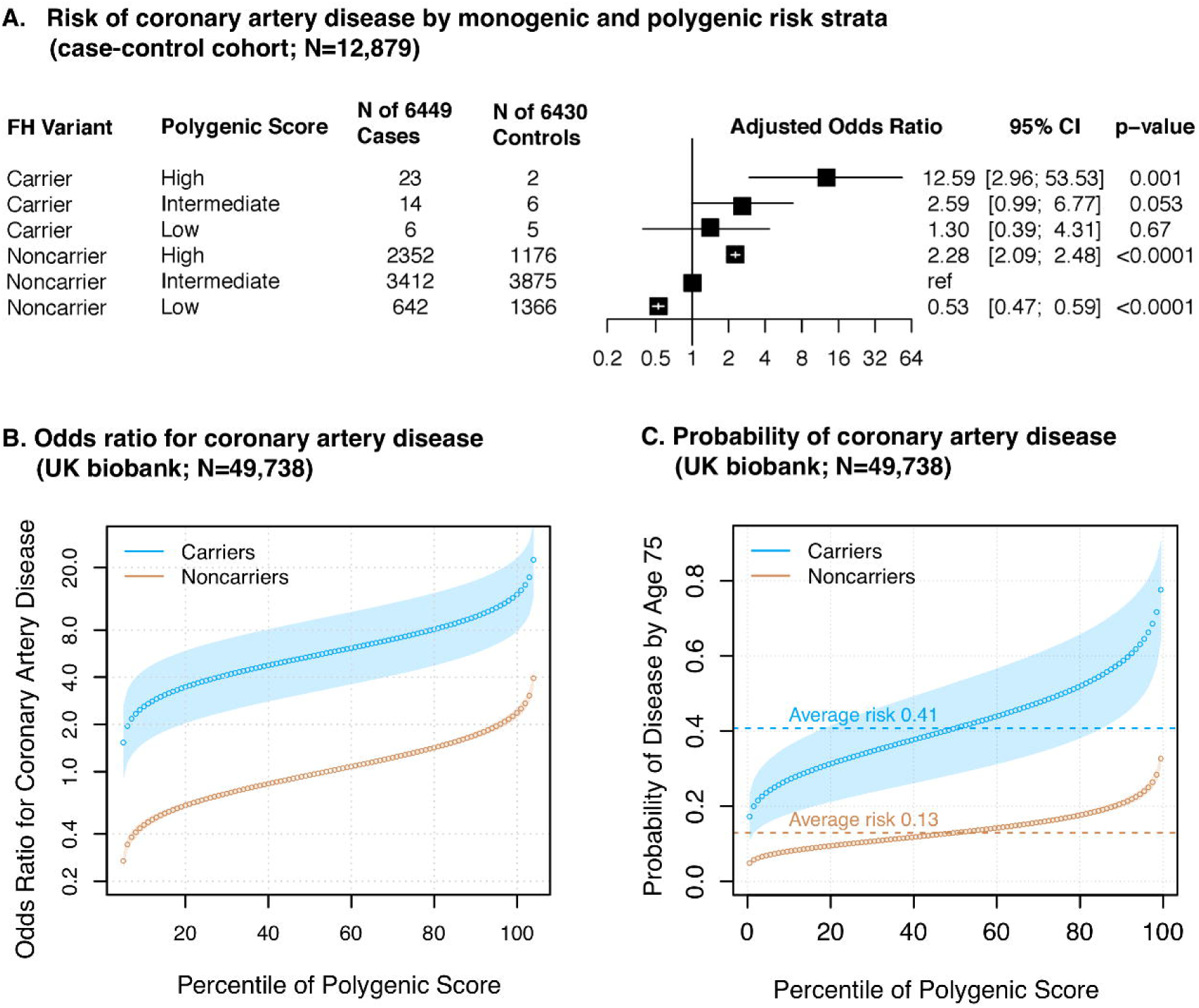
Polygenic background and risk of coronary artery disease in familial hypercholesterolemia. In panel A, case-control participants for coronary artery disease in the UK Biobank (n=12,879) were stratified into three groups according to their polygenic score — low, intermediate, or high defined as the lowest quintile, the middle three quintiles, and the highest quintile of the polygenic score distribution respectively. For carriers and noncarriers of familial hypercholesterolemia in each polygenic score group, the odds ratio for coronary artery disease was calculated in a logistic regression model with age, sex, and the first four principal components of ancestry as covariates. Non-carriers with intermediate polygenic score served as the reference group. In panels B and C, for UK Biobank participants (n=49,738), shown are the predicted odds ratios of coronary artery disease (panel B) and predicted probability of coronary artery disease by age 75 years (panel C) in each percentile (dots) of the polygenic score distribution for carriers (blue) and noncarriers (brown) of familial hypercholesterolemia variants. For calculating the odds ratios, a logistic regression model with age, sex, and the first four principal components of ancestry was used, and the predicted odds ratios were calculated by referencing to the risk of average polygenic risk score and conditioning on the mean value of each covariate. The predicted probability of disease as a function of monogenic variant carrier status and polygenic score was modeled using Cox regression models with age, sex, and the first four principal components of ancestry as covariates. Prevalent cases at baseline enrollment in the UK Biobank and incident cases during follow-up were included (Table S2 in the Supplementary Appendix). The shaded area around the dots represents the 95% confidence interval. The horizontal dashed lines show the odds ratio or probability of disease for people with average polygenic risk score. FH — familial hypercholesterolemia

We next examined an independent cohort of 49,738 UK Biobank participants, after confirming that monogenic variants and the polygenic score were both associated with coronary artery disease as expected (Tables S1-2 in the Supplementary Appendix). A laboratory geneticist identified a familial hypercholesterolemia variant in 131 (0.26%) of these participants, which conferred a 5.02-fold (95%CI 2.97 to 8.46) increased risk of coronary artery disease at the time of enrollment. The polygenic score for coronary artery disease was normally distributed in the population and strongly associated with disease — odds ratio per standard deviation increment of 1.64 (95%CI 1.57 to 1.72). Within a logistic regression model, the relationship between the polygenic score and prevalent disease conformed to a linear model (Fig. S2 and Table S3 in the Supplementary Appendix).

Joint modeling of monogenic variant carrier status and polygenic score indicate substantial gradients in risk of coronary artery disease according to inherited DNA variation that can be assessed from the time of birth. Odds ratio for coronary artery disease in monogenic variant carriers — as compared to noncarriers with average polygenic score — ranged from 1.59 to 21.24 across percentiles of the polygenic score (Figure 1B). Modeling the probability of disease by age 75 years indicated striking gradients in risk, ranging from 4.8% for individuals who were both noncarriers and in the lowest percentile of the polygenic score to 77.7% for individuals with a monogenic risk variant who were also in the highest polygenic score percentile (Figure 1C).

### Polygenic background modifies risk of breast cancer conferred by familial hereditary breast and ovarian cancer variants

We next set out to apply the same analysis to breast cancer. We first identified monogenic risk variants by sequencing the *BRCA1* and *BRCA2* genes in 1920 breast cancer cases and 17,344 controls, all female, from the Color Genomics commercial testing laboratory (Table 2). A pathogenic or likely pathogenic variant was identified in 176 (9.1%) cases and 674 (3.9%) controls, corresponding to a 3.48-fold (95% CI 2.81 to 4.21) increased risk of breast cancer in variant carriers. We then calculated polygenic risk for breast cancer, using a previously validated polygenic score. As we saw for coronary artery disease, breast cancer risk was strongly affected by polygenic background. Compared to noncarriers with intermediate polygenic score, increased risk among carriers ranged from 2.40-fold (95% CI 1.58 to 3.65) for those in the lowest quintile of the polygenic score distribution to 6.85-fold (95% CI 4.71 to 9.96) in the highest quintile (Figure 2B).

**Table 2:**
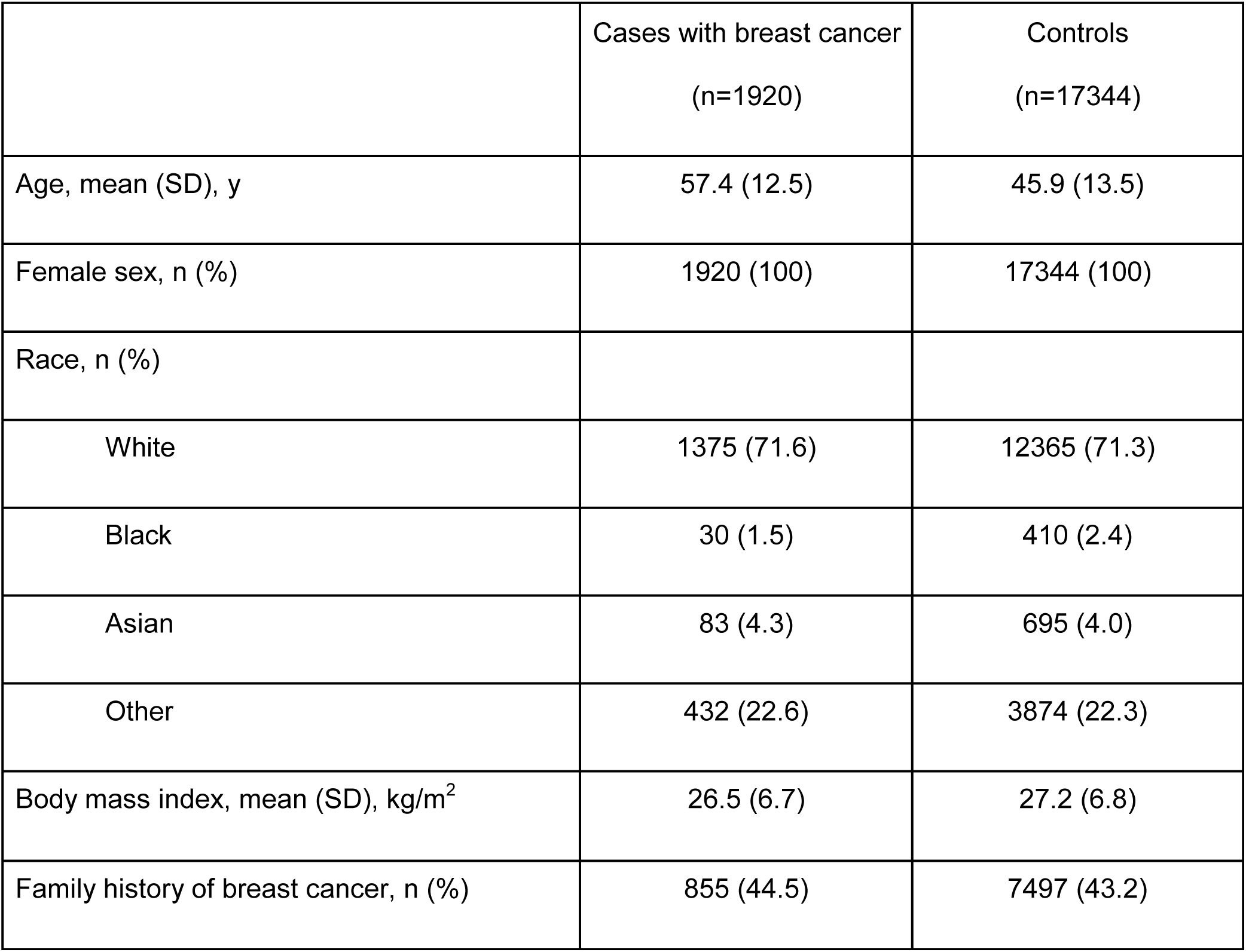
Baseline characteristics of breast cancer case-control study participants derived from the Color Genomics commercial testing laboratory

**Figure 2:**
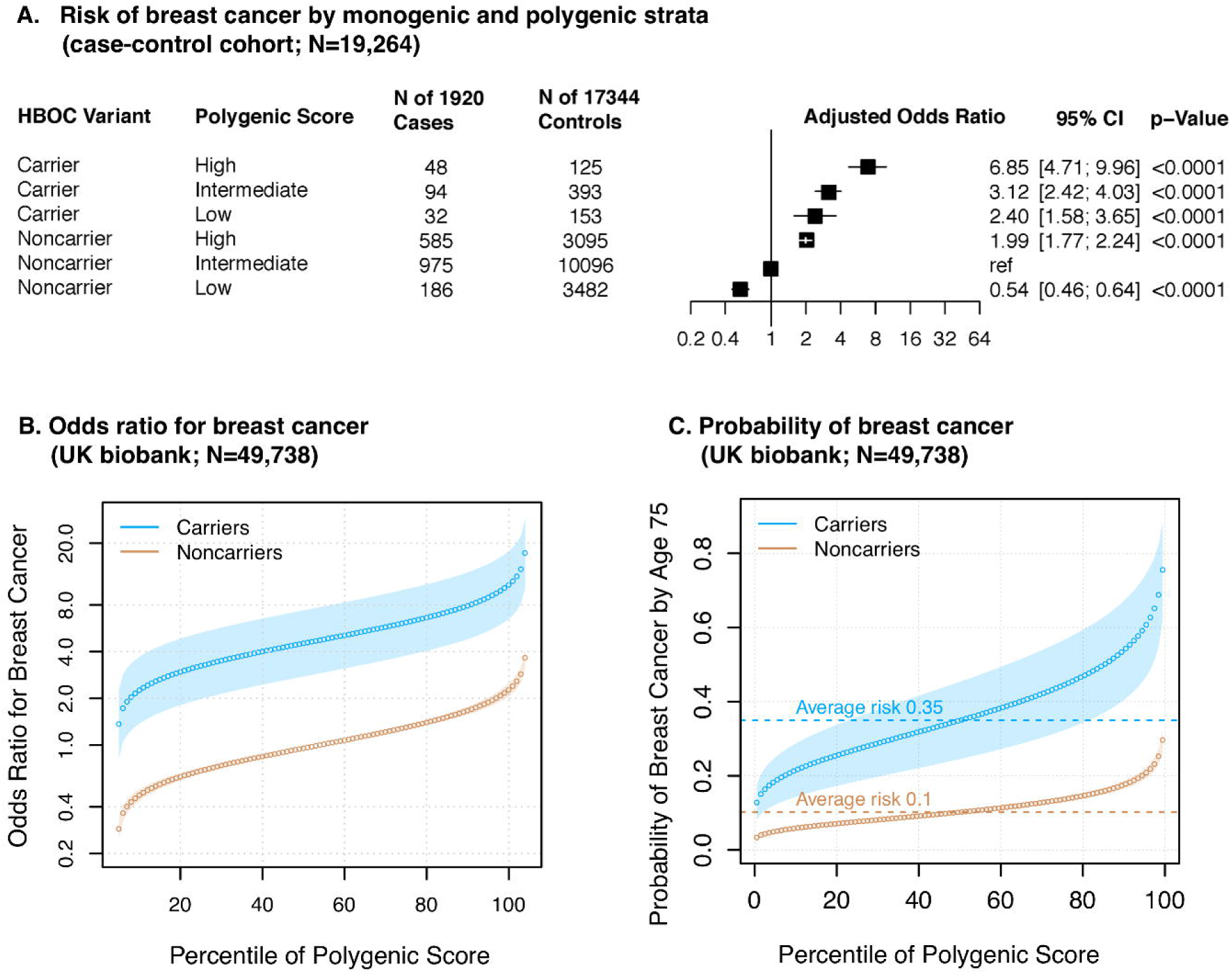
Polygenic background and risk of breast cancer in hereditary breast and ovarian cancer. In panel A, case-control participants for breast cancer in Color Genomics (n=19,264) were stratified into three groups according to their polygenic score — low, intermediate, or high defined as the lowest quintile, the middle three quintiles, and the highest quintile of the polygenic score distribution respectively. For carriers and noncarriers of hereditary breast and ovarian cancer variants in each polygenic score group, the odds ratio for breast cancer was calculated in a logistic regression model with age and the first four principal components of ancestry as covariates. Non-carriers with intermediate polygenic score served as the reference group. In panels B and C, for female UK Biobank participants (n=27,144), shown are the predicted odds ratios of breast cancer (panel B) and predicted probability of breast cancer by age 75 years (panel C) in each percentile (dots) of the polygenic score distribution for carriers (blue) and noncarriers (brown) of hereditary breast and ovarian cancer variants. For calculating the odds ratios, a logistic regression model with age and the first four principal components of ancestry was used, and the predicted odds ratios were calculated by referencing to the risk of average polygenic risk score and conditioning on the mean value of each covariate. The predicted probability of disease as a function of monogenic variant carrier status and polygenic score was modeled using Cox regression models with age and the first four principal components of ancestry as covariates. Prevalent cases at baseline enrollment in the UK Biobank and incident cases during follow-up were included (Table S2 in the Supplementary Appendix). The shaded area around the dots represents the 95% confidence interval. The horizontal dashed lines show the odds ratio or probability of disease for people with average polygenic risk score. HBOC — hereditary breast and ovarian cancer

We extended these results into the UK Biobank participants, this time focusing only on the 27,144 female participants. A laboratory geneticist reviewed all observed genetic variants in the *BRCA1* and *BRCA2* genes, identifying 116 carriers of pathogenic or likely pathogenic variants. These variants conferred a 4.45-fold increased risk of breast cancer (95%CI 2.75 to 7.20). The polygenic score was associated with an odds ratio per standard deviation increment of 1.61 (95%CI 1.52 to 1.70), with the relationship to breast cancer again conforming to a linear model (Fig. S2 and Table S3 in the Supplementary Appendix).

Joint modeling of both monogenic variant status and polygenic score indicated that the risk for breast cancer among carriers of a *BRCA1* or *BRCA2* variant ranges from 1.43 to 16.41 increased risk across percentile of the polygenic score. When modeled as probability of disease by age 75 years, risk among monogenic variant carriers ranged from 12.8 to 75.5% and risk among noncarriers ranged from 3.4 to 29.7%.

### Risk of colorectal cancer varies according to Lynch syndrome monogenic variant carrier status and polygenic score

We explored a third disease, colorectal cancer, in the same set of 49,738 UK Biobank participants used above. A pathogenic or likely pathogenic Lynch syndrome variant was identified in 76 (0.15%) individuals, conferring an odds ratio for colorectal cancer of 27.69 (95%CI 14.27 to 53.72). The odds ratio per standard deviation increment in the colorectal cancer polygenic score was 1.66 (95%CI 1.48 to 1.85). Joint modeling of monogenic variants and the polygenic score — using noncarriers with average polygenic score as the reference group — noted odds ratios ranging from 8.34 to 117.53 for carriers of monogenic variants and 0.27 to 3.78 for noncarriers (Figure 3A). Absolute risk of colorectal cancer by age 75 years ranged from 11.6 to 79.5% for carriers and 0.7 to 8.7% for noncarriers (Figure 3B).

**Figure 3:**
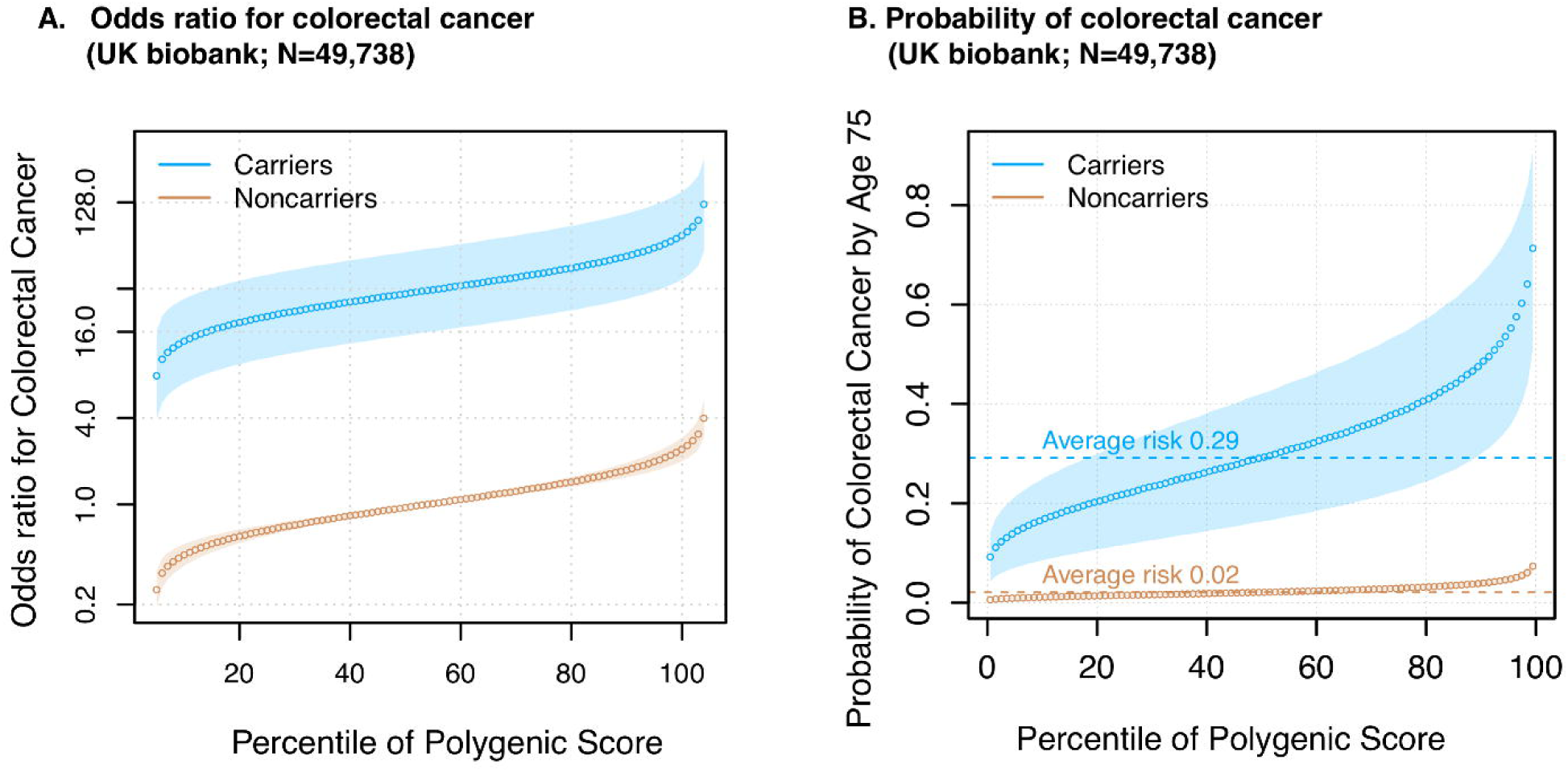
Polygenic background and risk of colorectal cancer in Lynch syndrome. For UK Biobank participants (n=49,738), shown are the predicted odds ratios of colorectal cancer (panel A) and predicted probability of colorectal cancer by age 75 years (panel B) in each percentile (dots) of the polygenic score distribution for carriers (blue) and noncarriers (brown) of Lynch syndrome variants. For calculating the odds ratios, a logistic regression model with age, sex, and the first four principal components of ancestry was used, and the predicted odds ratios were calculated by referencing to the risk of average polygenic risk score and conditioning on the mean value of each covariate. The predicted probability of disease as a function of monogenic variant carrier status and polygenic score was modeled using Cox regression models with age, sex, and the first four principal components of ancestry as covariates. Prevalent cases at baseline enrollment in the UK Biobank and incident cases during follow-up were included (Table S2 in the Supplementary Appendix). The shaded area around the dots represents the 95% confidence interval. The horizontal dashed lines show the odds ratio or probability of disease for people with average polygenic risk score.

## Discussion

Our analysis of the interaction between monogenic risk variants and polygenic background — for three important genomic conditions: familial hypercholesterolemia, hereditary breast and ovarian cancer syndrome, and Lynch syndrome — has two important implications.

First, we were surprised that risk conferred by monogenic risk variants, which act by perturbing a specific molecular pathway, can be so substantially modified by polygenic background, which appears to act by affecting a diverse set of physiological processes. From a physiological standpoint, it is not clear why the major disruptions caused by monogenic variants can be offset by other factors. Yet, the risk for monogenic variant carriers with the lowest polygenic risk scores approached the population average. Understanding the physiological basis for these interactions may suggest therapeutic strategies for monogenic variant carriers in general.

Second, our findings indicate that accounting for polygenic background is likely to increase the accuracy of risk estimation for individuals who inherit a monogenic risk variant. An important example is the decision about whether to undergo prophylactic mastectomy (as opposed to serial imaging) faced by carriers of a *BRCA1* variant.^17^ At present, up to 50% of women opt for prophylactic mastectomy, but rates are highly variable.^18,19^ Here, we find a broad spectrum of risk across percentiles of the polygenic score that may better inform shared decision making — odds ratios compared to noncarriers ranging from 1.4 to 16.4 and an absolute risk at age 75 years ranging from 13 to 76%. Similarly, refined risk estimates may improve decisions about the timing and intensity of lipid-lowering therapy for individuals with familial hypercholesterolemia and whether to undergo serial colonoscopies or prophylactic colectomy for individuals with Lynch syndrome variants.

In a clinical setting, assessing both monogenic risk variants and polygenic risk requires high-coverage sequencing of genes associated with monogenic risk and an approach to assay common variants across the genome. At present, this can be accomplished by various approaches, including (i) high-coverage whole genome sequencing,^15^ (ii) whole exome sequencing and genotyping array, or (iii) targeted high-coverage sequencing of individuals genes and with low-coverage sequencing across the genome.^20^ Ongoing efforts to improve the cost and accessibility of these technologies will improve the feasibility of incorporating this information into routine clinical practice.

From a statistical standpoint, our data indicate that polygenic risk contributes to the incomplete penetrance of the monogenic risk variants. Additionally, our data suggest a roughly additive interaction between monogenic risk variants and polygenic risk, although the statistical power to detect non-additive interaction was limited by the number of individuals carrying monogenic risk variants. Within the cohort of 49,738 UK Biobank participants, the p-values for interaction were 0.07, 0.53, and 0.49 for coronary artery disease, breast cancer, and colorectal cancer, respectively.

Our results should be interpreted in the context of potential limitations. First, UK Biobank participants tend to be healthier than the general population. Disease risk models should be calibrated for a given target population prior to clinical use.^21^ Second, our analysis focused only on the role of the monogenic variants for the three conditions studied; for example, it did not consider the risk of the breast and ovarian cancer variants on ovarian cancer.^22^ Third, we aggregated together all pathogenic and likely pathogenic variants for each monogenic condition; however; there may be heterogeneity among these variants.^22–24^ Fourth, further efforts are needed to understand how best to disclose integrated genomic risk assessments to patients and treating clinicians and how to integrate them with existing predictors based on nongenetic factors.^25–27^

Finally, we highlight an important equity issue. The fact that our knowledge concerning the monogenic risk variants and the development of the polygenic scores has been based primarily on patients of European ancestry affects the utility for patients of other ancestries.^15,28–30^ In particular, the polygenic scores are known to be less precise for other ancestry groups.^30^ It is important for the biomedical community to invest in the development of more diverse population allele frequency databases,^31^ disease association studies in other ancestral backgrounds, new computational algorithms that better account for ancestral background,^32^ and new technology or machine learning algorithms to enable unbiased high-throughput functional assessments of variants.^33,34^

## Data Availability

Data on UK biobank participants is available to approved investigators directly form the UK biobank.

## Notes

### Competing Interest Statement

A.C.F. is a consultant and holds equity in GoodPath. A.P. is a Venture Partner at GV, a subsidiary of Alphabet Corporation. P.T.E. is supported by a grant from Bayer AG to the Broad Institute focused on the genetics and therapeutics of cardiovascular diseases, and has served on advisory boards or consulted for Bayer AG, Quest Diagnostics, and Novartis. S.K. is an employee of Verve Therapeutics, and holds equity in Verve Therapeutics, Maze Therapeutics, Catabasis, and San Therapeutics. He is a member of the scientific advisory boards for Regeneron Genetics Center and Corvidia Therapeutics; he has served as a consultant for Acceleron, Eli Lilly, Novartis, Merck, Novo Nordisk, Novo Ventures, Ionis, Alnylam, Aegerion, Haug Partners, Noble Insights, Leerink Partners, Bayer Healthcare, Illumina, Color Genomics, MedGenome, Quest, and Medscape; he reports patents related to a method of identifying and treating a person having a predisposition to or afflicted with cardiometabolic disease (20180010185) and a genetics risk predictor (20190017119). K.N. is an employee of IBM Research. A.V.K. has served as a consultant or received honoraria from Novartis Institute for Biomedical Research, Amarin Pharmaceuticals, Maze Therapeutics, Color Genomics, Illumina, and Navitor Pharmaceuticals, received grant support from the Novartis Institute for Biomedical Research and IBM Research, and reports a patent related to a genetic risk predictor (20190017119). J.R.H, C.L.N., C.L., and A.Y.Z. are employees of Color Genomics. E.S.L. serves on the Board of Directors for Codiak BioSciences and Neon Therapeutics; serves on the Scientific Advisory Board of F-Prime Capital Partners and Third Rock Ventures; serves on the Board of Directors of the Innocence Project, Count Me In, and Biden Cancer Initiative; and serves on the Board of Trustees for the Parker Institute for Cancer Immunotherapy. The remaining authors have no disclosures.

### Funding Statement

Funding support was provided by grant T32HL007208 from the National Heart, Lung, and Blood Institute (A.P.P. and A.C.F.), grant 14CVD01 Fondation Leducq (P.T.E.), grants 1RO1HL092577, R01HL128914, K24HL105780 from the National Heart, Lung, and Blood Institute (P.T.E), grant 18SFRN34110082 from the American Heart Association (P.T.E.), an institutional grant from the Broad Institute of MIT and Harvard (BroadIgnite, to A.V.K.), grant 1K08HG010155 (to A.V.K.) and R01HG010372 (to C.A.C. and M.L.) from the National Human Genome Research Institute, a Hassenfeld Scholar Award from Massachusetts General Hospital (to A.V.K.), and a sponsored research agreement from IBM Research (to A.P., A.V.K.).

